# Evidence for trans-synaptic propagation of oligomeric tau in Progressive Supranuclear Palsy

**DOI:** 10.1101/2022.09.20.22280086

**Authors:** Robert I McGeachan, Lois Keavey, Jamie L Rose, Elizabeth M Simzer, Ya Yin Chang, Maxwell P Spires-Jones, Mollie Gilmore, Natalia Ravingerova, Cristina Scutariu, Lewis Taylor, Declan King, Makis Tzioras, Jane Tulloch, Sam A Booker, Imran Liaquat, Nicole Hindley-Pollock, Bethany Geary, Colin Smith, Paul M Brennan, Claire S Durrant, Tara L Spires-Jones

**Author notes:** Correspondence to, Prof Tara Spires-Jones Or, Dr Claire Durrant, Centre for Discovery Brain Sciences The University of Edinburgh, 1 George Square, Edinburgh EH8 9JZ, UK. These authors contributed equally (Claire S Durrant, Tara L Spires-Jones).

## Abstract

In the neurodegenerative disease Progressive Supranuclear Palsy (PSP), tau pathology progresses through the brain in a stereotypical spatiotemporal pattern, and where tau pathology appears, synapses are lost. We tested the hypothesis that tau pathology spreads between brain regions in PSP by moving from pre- to post-synapses. Sub-diffraction-limit microscopy of human post-mortem brain samples revealed that oligomeric tau is present in synaptic pairs in PSP, with an 80-fold increased chance of post-synapses containing tau when they oppose a tau-containing pre-synapse. In living human brain slice cultures, PSP-derived oligomeric tau was taken up by post-synapses. Synaptic engulfment by astrocytes was observed in both post-mortem brain and human brain slice cultures challenged with PSP-derived tau. These data indicate that tau pathology spreads via synapses in PSP and that astrocytes contribute to synapse loss. Targeting synaptic tau and astrocyte-mediated phagocytosis of synapses are promising targets for attenuating synaptic loss and pathology propagation in PSP.

## Introduction

Progressive Supranuclear Palsy (PSP) is a neurodegenerative disease characterized by accumulation of tau pathology in neurons, oligodendrocytes, and astrocytes, accompanied by impairments in movement, balance, cognition, and behaviour(1). Tau pathology accumulates in the brain in a stereotypical spatiotemporal pattern as PSP progresses (2). Areas with a high tau burden show synaptic loss and the degree of synapse loss over time correlates with the progression of disease symptoms (3,4). Tau protein can bind synaptic vesicles and affects synaptic function (5), and pathological phosphorylation and aggregation of tau cause synaptic dysfunction and loss in model systems (6–10). Thus, understanding the mechanisms of tau propagation and how pathological tau causes synapse loss are important for future therapeutic development to slow or stop symptoms of PSP.

The mechanisms driving tau propagation and synaptic loss in PSP remain largely unknown. Although one hypothesis is that the stereotypical tau accumulation in sequential brain regions in tauopathies is due to different regions being differentially vulnerable to local pathological changes in tau (11), there is strong evidence suggesting that pathologically aggregated and phosphorylated tau spreads physically through the brain both within local regions and between different brain regions. In mouse models of tauopathy, tau pathology can spread between brain regions by being released from presynaptic terminals and taken up by connected post-synapses. For example, in mice expressing mutant human tau with expression restricted to neurons in the entorhinal cortex, tau pathology spreads via the perforant pathway to connected dentate gyrus neurons (which do not express human tau) via pre-to-post synapse spread (12–17). More specific to PSP, injecting extracts of post-mortem PSP brain into mice expressing wild-type human tau induces formation of neuronal and glial inclusions which spread through the brain (18), and recent research suggests that the progression of tau pathology in human PSP occurs along functionally connected brain circuits (19).

Recently our group used array tomography and electron microscopy to study synaptic accumulation and potential trans-synaptic propagation of tau in Alzheimer’s disease, the most common tauopathy. We observed that oligomeric tau accumulates in synaptic pairs, supporting the potential spread between synaptically-connected neurons (20). Further, oligomeric tau was observed in presynaptic terminals in the occipital cortex, one of the last brain areas in Alzheimer’s disease to accumulate tau pathology. This supports the hypothesis that presynaptic release of oligomers from affected brain regions is a mechanism of pathological tau spread (20). Although fibrillar tau deposits, such neurofibrillary tangles (NFTs) and tufted astrocytes (TA) are the pathological hallmarks of tauopathies, research in model systems suggests that these are likely end-stage aggregates and do not themselves induce neurodegeneration or functional abnormalities (21–23). There is growing body of evidence that suggests that soluble tau oligomers are the source of toxicity, instead of large pathological aggregates including NFTs. Soluble tau oligomers induce early synaptic dysfunction, synaptic and neuronal degeneration, and have a greater propensity for propagation and seeding than fibrils (24–26). One potential mechanism of synaptotoxicity is through oligomeric tau binding synaptic vesicles and impairing neurotransmitter release. In post-mortem tissue from individuals with Alzheimer’s disease and in *Drosophila* and mouse models, tau colocalises with the presynaptic vesicle protein synaptogyrin-3 (9,10,27). In the animal models, lowering synaptogyrin-3 levels prevented synapse loss and memory decline (9,10,27). This indicates that disrupting this tau-synaptogyrin-3 interaction is a promising therapeutic avenue to prevent synaptic dysfunction, loss, and downstream cognitive deficits.

Another potential mechanism leading from pathological tau to synapse loss is via aberrant induction of synaptic pruning by glia. Synaptic pruning mediated by astrocytes and microglia is an important aspect of normal circuit development (28–31). Recently, we observed astrocytes and microglia engulf synapses in Alzheimer’s disease, with astrocytes containing more synaptic proteins than microglia (32). Further, in AD, presynapses containing p-tau Ser356 are five times more likely to colocalise with astrocytes (33). Similarly, astrocytes and microglia contain synaptic tau synapses containing tau oligomers are more likely to be engulfed by glia (34). In AD, amyloid pathology is thought to drive a large amount of the observed neuroinflammation. In our study for example, glial engulfment of synapses was greatest in the vicinity of amyloid plaques (32). Less is known about glial engulfment of synapses in primary tauopathies like PSP despite prominent astrocyte pathology.

While synapse loss and the spread of tau pathology have been observed in PSP as detailed above, there have not previously been detailed investigations of synaptic tau or potential mechanisms of tau spread and toxicity in human PSP. Using post-mortem PSP brain and living human brain slice cultures, here we test the hypothesis that tau pathology may spread trans-synaptically in PSP and that synaptic accumulation of tau contributes to synapse loss by binding synaptogyrin-3 and inducing synaptic engulfment by astrocytes.

## Results

### Phosphorylated tau colocalises with pre-synaptic vesicle proteins in PSP

To examine sub-synaptic localisation of tau, post-mortem brain tissue from substantia nigra and frontal cortex of people who died with PSP and control donors was prepared for array tomography (details of donors are found in table 1). Ribbons of serial ultrathin sections were immunostained for phospho-tau Ser202/Thr205 (AT8) and synaptophysin (Fig. 1a). Analysis of the proportion of synapses containing tau staining was performed and linear mixed effect modelling of Tukey transformed data (%_synapses_with_tau ∼ Diagnosis*Brain_region 1|Case_ID). We observe an accumulation of AT8-positive phospho-tau in synapses with a higher proportion in PSP brains than in controls (median 0.25% of presynaptic terminals contain phospho-tau in PSP, 0% in controls, ANOVA after linear mixed effects model F[1,21]=89.24, *p*<0.0001). Post-hoc analyses show this increase in colocalization of AT8 and synaptophysin in PSP is significant in both in both the substantia nigra (β = 0.787, 95% CI [0.565, 1.009], *t*(21) = 7.377, *p* < 0.0001) and frontal cortex (β = 0.557, 95% CI [0.361, 0.753], *t*(21) = 5.918, *p* < 0.0001) (Fig. 1b). Within PSP cases there was more AT8 positive tau in synapses in the substania nigra compared to the frontal cortex (β = 0.2705, 95% CI [0.056, 0.485], *t*(21) = 2.622, *p* = 0.016)(Fig. 1b).

**Figure 1:**
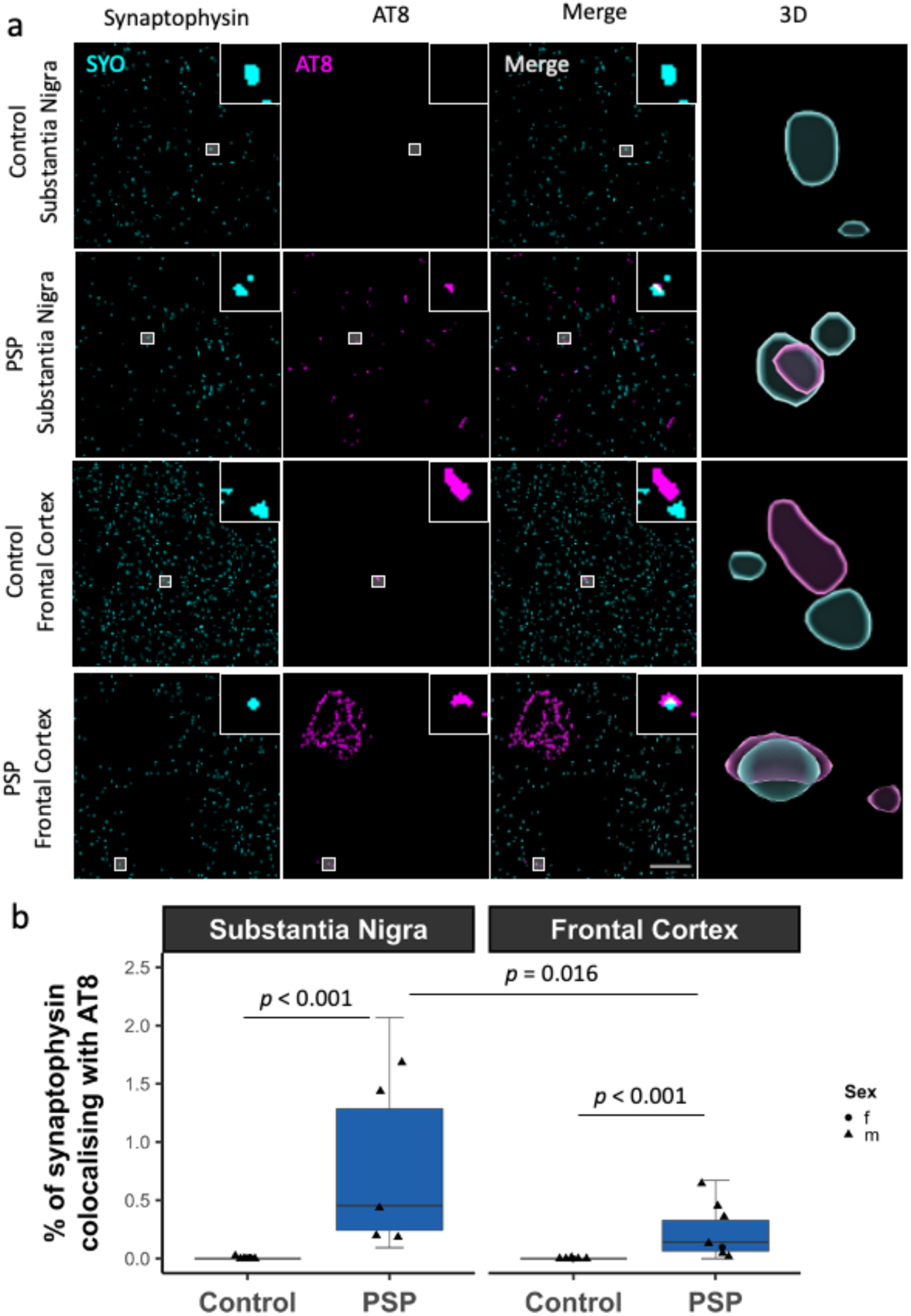
Phospho-tau accumulates within in pre-synaptic terminals in PSP. **a)** Array tomography ribbons were immunostained for synaptophysin (cyan) and phospho-tau Thr202, Ser205 (AT8, magenta). Single 70nm segmented sections show representative staining in the substantia nigra and frontal cortex of control and PSP brain. Inserts highlight colocalization between synaptophysin and AT8. 3D reconstructions made using IMARIS. Scale bar = 10 μm. Large boxes = 50 μm * 50 μm. Inserts = 2 μm * 2 μm. **b)** Quantification of colocalisation shows an increase in the percentage of synaptophysin puncta colocalizing with AT8 in PSP brain compared to control. Within PSP brain, there are more synaptophysin objects colocalizing with AT8 in the substantia nigra compared to the frontal cortex. Boxplots show quartiles and medians calculated from each image stack. Data points refer to case means (females, circles; males, triangles).

**Table 1.** Details of cases used in human post-mortem experiments. 1 = Array tomography on frontal cortex (fig 1, 2, 3, 4); 2 = Array tomography on substantia nigra (fig 1, 4); 3 = Proteomics analysis of frontal cortex (fig 6); 4 = Proteomics analysis of substantia nigra (fig 6). BBN = UK Brain Bank Number.

| Diagnosis | BBN | SD Number | Age | Sex | PMI | Brain Weight | Brain pH | Experiments |
| --- | --- | --- | --- | --- | --- | --- | --- | --- |
| PSP | 1.32816 | SD006/18 | 70s | M | 114 | 1309 | 6 | 1,2,3,4 |
| PSP | 1.35381 | SD009/19 | 70s | M | 66 | 1242 | 6.19 | 1,2,3,4 |
| PSP | 1.30843 | SD030/17 | 70s | M | 46 | 1477 | 6.12 | 1,2,3,4 |
| PSP | 1.30855 | SD033/17 | 80s | M | 89 | 1197 | 6.15 | 1,2,3,4 |
| PSP | 1.32508 | SD049/17 | 60s | M | 79 | 1325 | 6 | 1,2,3,4 |
| PSP | 1.36418 | SD008/21 | 60s | F | 62 | 1036 | 6.21 | 1,3,4 |
| PSP | 1.36927 | SD041/21 | 70s | M | 103 | 1457 | 5.95 | 1,3,4 |
| PSP | 1.34194 | RU61/201<br>8 | 80s | M | 144 | N/A | N/A | 3 |
| PSP | 1.37236 | SD012/22 | 80s | M | 78 | 1155 | 6.59 | 3,4 |
| PSP | 1.35439 | SD018/19 | 70s | F | 91 | 1328 | 5.72 | 3,4 |
| Control | 1.29880 | SD010/17 | 50s | M | 64 | 1520 | 6.41 | 1,2,3,4 |
| Control | 1.28794 | SD017/16 | 70s | F | 72 | 1219 | 5.95 | 1,2,3,4 |
| Control | 1.26495 | SD024/15 | 70s | M | 39 | 1290 | 6.17 | 1,2,3,4 |
| Control | 1.28797 | SD025/16 | 70s | M | 57 | 1301 | 6.11 | 1,2,3,4 |
| Control | 1.29084 | SD032/16 | 50s | M | 104 | 1560 | 6.14 | 1,2,3,4 |
| Control | 1.28402 | SD051/15 | 70s | M | 49 | 1503 | 6.33 | 1,2,3,4 |
| Control | 1.35529 | SD026/19 | 50s | M | 96 | 1650 | 6.49 | 1,3,4 |
| Control | 1.32848 | SD011/18 | 60s | F | 80 | 1323 | 5.77 | 3 |
| Control | 1.31504 | SD046/17 | 60s | F | 76 | 1150 | 6.35 | 3 |
| Control | 1.28406 | SD001/16 | 70s | M | 72 | 1437 | 6.13 | 3,4 |
| Control | 1.29525 | SD042/16 | 50s | M | 52 | 1500 | 6.3 | 3,4 |
| Control | 1.28794 | SD018/16 | 70s | F | 61 | 1289 | 5.76 | 4 |
| Control | 1.19686 | SD063/13 | 70s | F | 75 | 1320 | 6.5 | 4 |

To test whether tau binds synaptogyrin-3 in human synapses as has been previously observed in mice and flies (10,27), ribbons from the frontal cortex were also immunostained for tau, synaptogyrin-3 and synaptophysin (Fig. 2a,b). Linear mixed effects model of Tukey transformed data with diagnosis as the dependent variable and case as a random effect demonstrated a 5-fold increase in colocalisation between total tau and synaptogyrin-3 in PSP (Fig 2c, median 0.16% of synapses in PSP, 0.03% in controls, β = 0.279, 95% CI [0.004, 0.553], *t*(11.68)=2.25, *p*=0.044). Similarly, the percentage of synaptophysin puncta containing total tau was significantly higher in PSP cases (Fig. 2d, median 0.04% in PSP, 0% in controls, β = 0.206, 95% CI [0.00004, 0.413] *t*(11.13)=2.24, *p*=0.047. As expected, the burden of tau pathology assessed as the percentage volume occupied by total tau staining was significantly higher in PSP cases (Fig. 2e, β = 0.427, 95% CI [0.251, 0.603], t=5.73, p<0.0001). Spearman’s correlation reveals that the proportion of synaptogyrin-3 puncta containing tau positively correlated with the tau burden in PSP cases (Fig. 2f, correlation coefficient = 0.93, p=0.007) but not in control cases (correlation coefficient=0.78, p=0.22).

**Figure 2:**
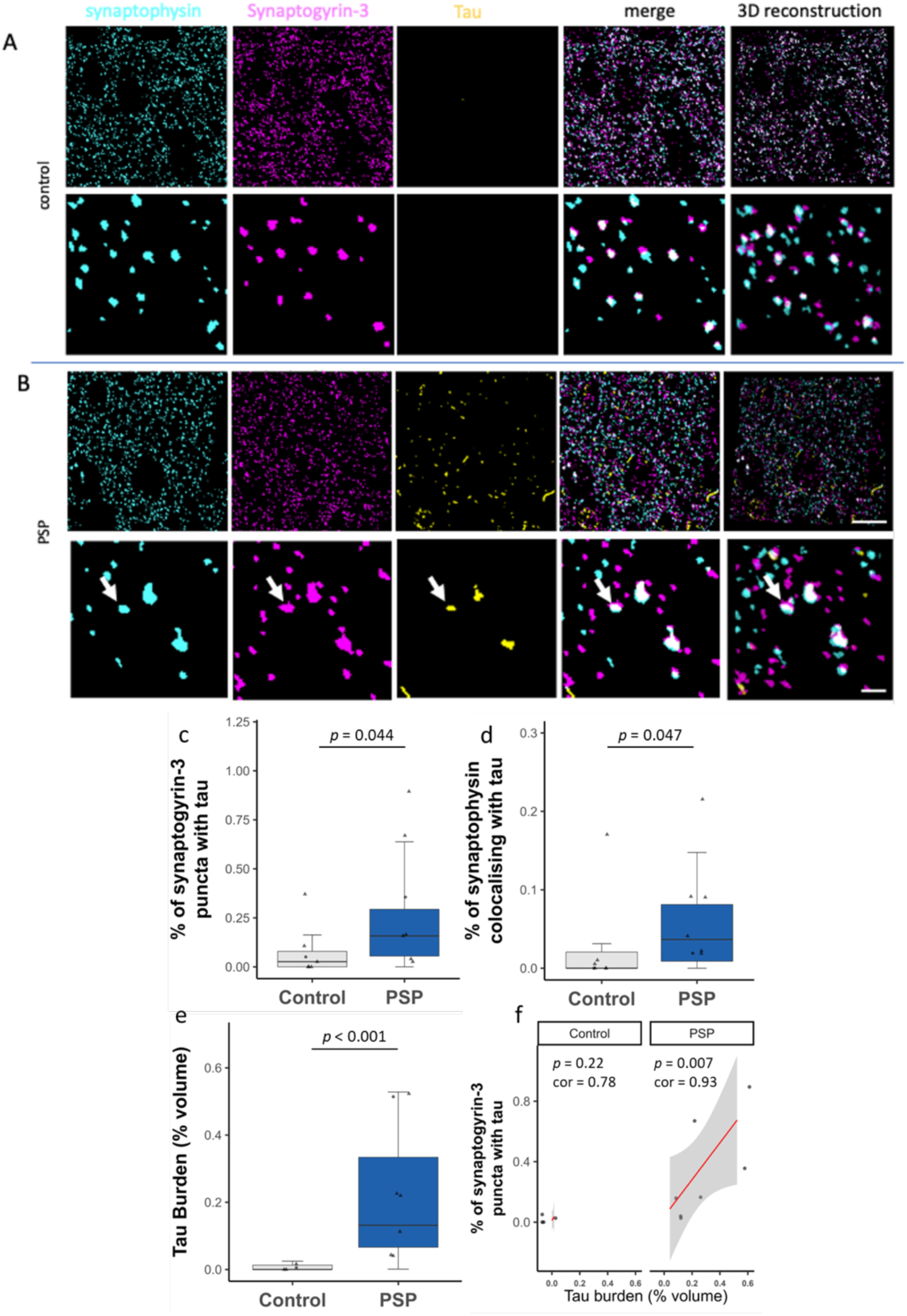
Synaptic tau colocalises with synaptogyrin-3 in PSP. **a-b)** segmented array tomography images. Array tomography ribbons were stained for synaptophysin (cyan), synaptogyrin-3 (magenta) and tau (stained with a polyclonal antibody to human tau, yellow). Representative images of maximum intensity z-projections of 5 serial sections are shown for a control (a) and a PSP case (b). The top row of each panel contain a 50x50μm region of interest with a 10μm scale bar and the bottom rows contain a zoomed in 10x10μm region of interest with a 2μm scale bar. The far-right column shows three-dimensional reconstructions of 5 serial sections. An arrow indicates a synapse containing tau in the PSP case. **c-f)** Quantification of segmented array tomography images reveals increases in the percentage of synaptogyrin-3 puncta containing tau (c), synaptophysin puncta containing tau (d), and increased tau burden (e). Tau burden positively correlates with the percentage of synaptogyrin-3 puncta containing tau in PSP but not control cases (f). Boxplots show quartiles and medians calculated from each image stack. Data points refer to case means (females, circles; males, triangles)

### Oligomeric tau accumulates within synaptic pairs in PSP: Indirect evidence for tau spread

We further used array tomography to investigate the synaptic distribution of oligomeric tau (T22 antibody, validated by Lasagna-Reeves (35)), in the frontal cortex. The frontal cortex was chosen for this experiment as it is a brain region that is later affected by tau pathology. Our reasoning for this was that, even though tau burden is lower in the frontal cortex, if trans-synaptic tau spread is a mechanism driving tau pathology progression, then a greater proportion of the tau present in the frontal cortex may have spread there, compared to an early affected region where disease initiation occurs. Array tomography ribbons were stained for oligomeric tau (T22), the pre-synaptic vesicle protein synaptophysin (SYO) and postsynaptic density protein 95 (PSD-95) (Fig. 3a). Colocalisation analyses were performed and linear mixed effect modelling of the data shows that, compared to control, in PSP there is an increased proportion of oligomeric tau containing pre-synapses (β = 0.429, 95% CI [0.234, 0.624], *t*(11) = 4.85, *p* = 0.0005) (Fig. 3b, e) and post-synapses (β = 0.606, 95% CI [0.332, 0.88], *t*(10.8) = 4.89, *p* = 0.0005)(Fig. 3c, f). Interestingly, there was a negative correlation between the percentage of oligomeric tau containing pre-(correlation coefficient = -0.597, *p* = 0.031)(Fig. 3h) and post-synapses (correlation coefficient = -0.579, *p* = 0.038)(Fig. 3i) and their respective synaptic density, indicating that oligomeric tau is synaptotoxic. Synaptic pairs were defined as a PSD95 puncta with a synaptophysin puncta within 0.5 μm (distance between centroids). This distance was chosen based on previous electron microscopy visualisation of synapses. In PSP brain we also observed an increase in synaptic pairs where tau oligomers were colocalising to both the pre- and opposing paired post-synapse (β = 0.3, 95% CI [0.077, 0.522], *t*(11.6) = 3.06, *p* = 0.010) (Fig. 3d, g). We then wanted to see how much more often tau accumulated in synaptic pairs than it would by chance, as evidence to support the theory of trans-synaptic tau spread. In PSP brain, we found that post-synapses paired to a pre-synapse containing oligomeric tau were 84-times more likely colocalise with tau, than all paired post-synapses regardless of the tau status of the paired pre-synapse (β = 27.7, 95% CI [6.54, 48.8], *t*(11.8) = 2.856, *p* = 0.015)(Fig. 3j). We found a similar finding when examining the presence of AT8 at synaptic pairs (supplementary fig. 1a-g).

**Figure 3:**
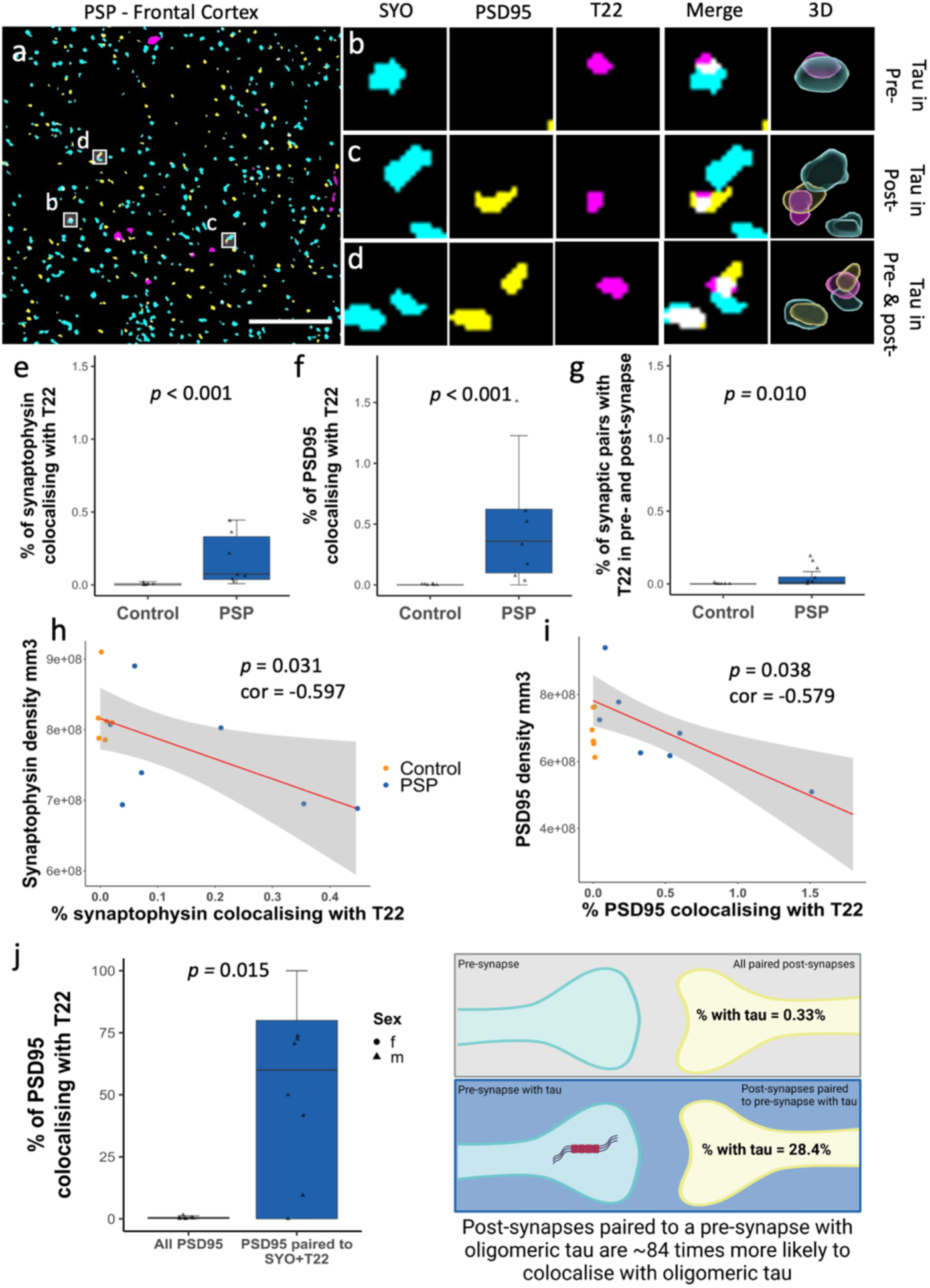
Indirect evidence for trans-synaptic spread of oligomeric tau. **a-d)** A single 70nm segmented section of PSP frontal cortex, immunostained for pre-synapses (synaptophysin, cyan), post-synapses (PSD95, yellow) and oligomeric tau (magenta). Scale bar = 10um. Oligomeric tau is seen in pre-synapses (b), post-synapses (c) and in both the pre- and post-synapse of a synaptic pair (d). Boxes = 2um * 2um, 3D reconstructions made using IMARIS. **e-j)** Quantification shows that in PSP frontal cortex there is an increase in the percentage of pre-synapse (e), post-synapses (f) and synaptic pairs (g) colocalising with oligomeric tau. The percentage of pre-synapses (h) and post-synapses (i) with oligomeric tau negatively correlates with the density of pre- and post-synapses respectively. Post-synapses that are paired to a pre-synapses with oligomeric tau are around 84 times more likely to colocalize with oligomeric tau than all paired post-synapses, regardless of whether the adjacent pre-synapse also colocalises with tau. Boxplots show quartiles and medians calculated from each image stack. Data points refer to case means (females, circles; males, triangles). Schematic created with biorender.com.

### Astrocytes show increased synaptic engulfment in PSP

To investigate the presence of astrogliosis and synaptic engulfment by astrocytes in PSP substantia nigra and frontal cortex, we immunostained array tomography ribbons for GFAP, synaptophysin and AT8 (Fig. 4a,b). We observe an increase in GFAP burden in both the substantia nigra (β = 0.541, 95% CI [-0.06, 1.14], *t*(20) = 2.206, *p* = 0.039) and frontal cortex (β = 0.139, 95% CI [-0.02, 0.30], *t*(20) = 2.22, *p* = 0.037)(Fig. 4c). There is also an increase in the percentage of pre-synapses colocalising with GFAP positive astrocytes in the substantia nigra (β = 0.374, 95% CI [0.04, 0.33], *t*(21) = 3.089, *p* = 0.006) and frontal cortex (β = 0.243, 95% CI [-0.01, 0.25], *t*(21) = 2.266, *p* = 0.034)(Fig. 4d). In some PSP cases, synaptophysin colocalised with both GFAP and AT8, suggesting that astrocytes might be engulfing tau containing synapses. In the frontal cortex, this analysis was repeated with PSD95 with comparable results (supplementary fig. 1c,h), suggesting that in the frontal cortex of PSP brain astrocytes are engulfing an increased number of both pre-synapses and excitatory post-synapses.

**Figure 4:**
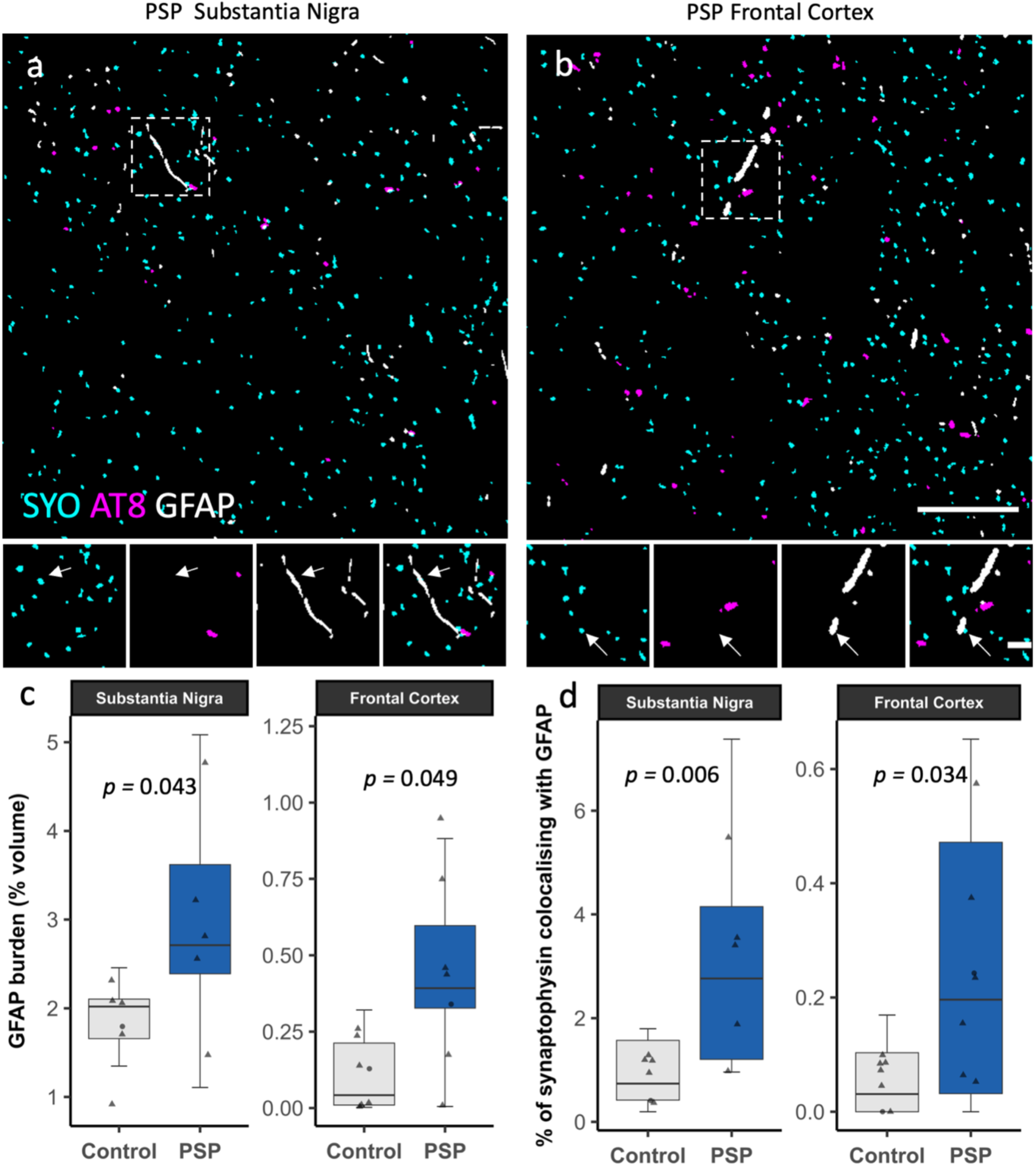
Astrogliosis and increased synaptic engulfment in PSP. **a-b)** A single 70nm segmented section of PSP substantia nigra (a) and frontal cortex (b), immunostained for pre-synapses (synaptophysin, cyan), tau (AT8, magenta) and astrocytes (GFAP, grey). Large boxes: 50μm*μm, scale bar = 50μm. Small boxes: 10μm *10μm, scale bar = 2μm. **c-d)** Quantification of segmented array tomography images reveals that in PSP substantia nigra and frontal cortex, there is an astrogliosis (c) and increased colocalisation between pre-synapses (synaptophysin) and astrocytes (GFAP) (d). Boxplots show quartiles and medians calculated from each image stack. Data points refer to case means (females, circles; males, triangles)

### Proteomics analysis of PSP post-mortem brain samples reveals dysregulated synaptic, inflammatory and metabolic pathways

Frozen samples from substantia nigra and frontal cortex of PSP and control cases were homogenized and synapses were enriched using a synaptoneurosome preparation. Proteomics was run on both total soluble homogenates and synaptic fractions. To confirm that our synaptic preparations were effective, we used western blots to confirm enrichment of synaptic proteins synaptophysin and PSD95 and exclusion of nuclear histone protein (supplementary fig. 2a-c). Further we confirmed predominantly synaptic structures using electron microscopy, and finally, Gene ontology (GO) analysis of cellular compartments present confirmed that our synaptoneurosome preparations were enriched for multiple synaptic proteins when compared to the total brain homogenate preparations (supplementary fig. 2e). The synaptic and total brain homogenate proteome were then compared between PSP and control frontal cortex and substantia nigra. Volcano plots illustrate the extent of protein changes detected between PSP and control (supplementary fig. 3). Differentially expressed proteins (DEPs) of interest, include the complement proteins C4A, C1QA, and C1QC, the neuroinflammatory proteins GFAP, YKL40/CHI3L1, and ICAM1, the synaptic protein synatxin-6 (STX6), stomatin and clusterin.

For a deeper understanding of the biological and synaptic pathways affected in PSP, DEPs were analysed utilizing GO enrichment (setting: biological process) (34) and the synaptic gene ontology tool, SynGO (35) (Fig. 5). Alterations were evident in several biological pathways in PSP, including changes in metabolic pathways such as mitochondrial translation and gene expression, oxidative phosphorylation, aerobic transport chain, and ATP synthesis. Alterations in immune and inflammatory pathways, such as complement activation, humoral immune response, detoxification, and stress response, were also observed. Dysregulation of synaptic and signalling pathways, such as neurotransmitter transport, neurotransmitter signalling, and vesicle fusion, was evident. Interestingly, while the same pathways were often altered in both the frontal cortex and substantia nigra, the direction of change was often reversed (Fig. 5a). This observation suggests a potential dichotomous response in end-stage and actively compensatory stages of the disease. To interrogate synaptic changes further, SynGO analysis was conducted on the synaptoneurosome preparations. In the substantia nigra, there was evidence for a loss of both pre- and post-synaptic proteins (Fig. 5b,c). In the frontal cortex, there was evidence for both a loss of some synaptic proteins and an increase in others (Fig. 5d,e).

**Figure 5:**
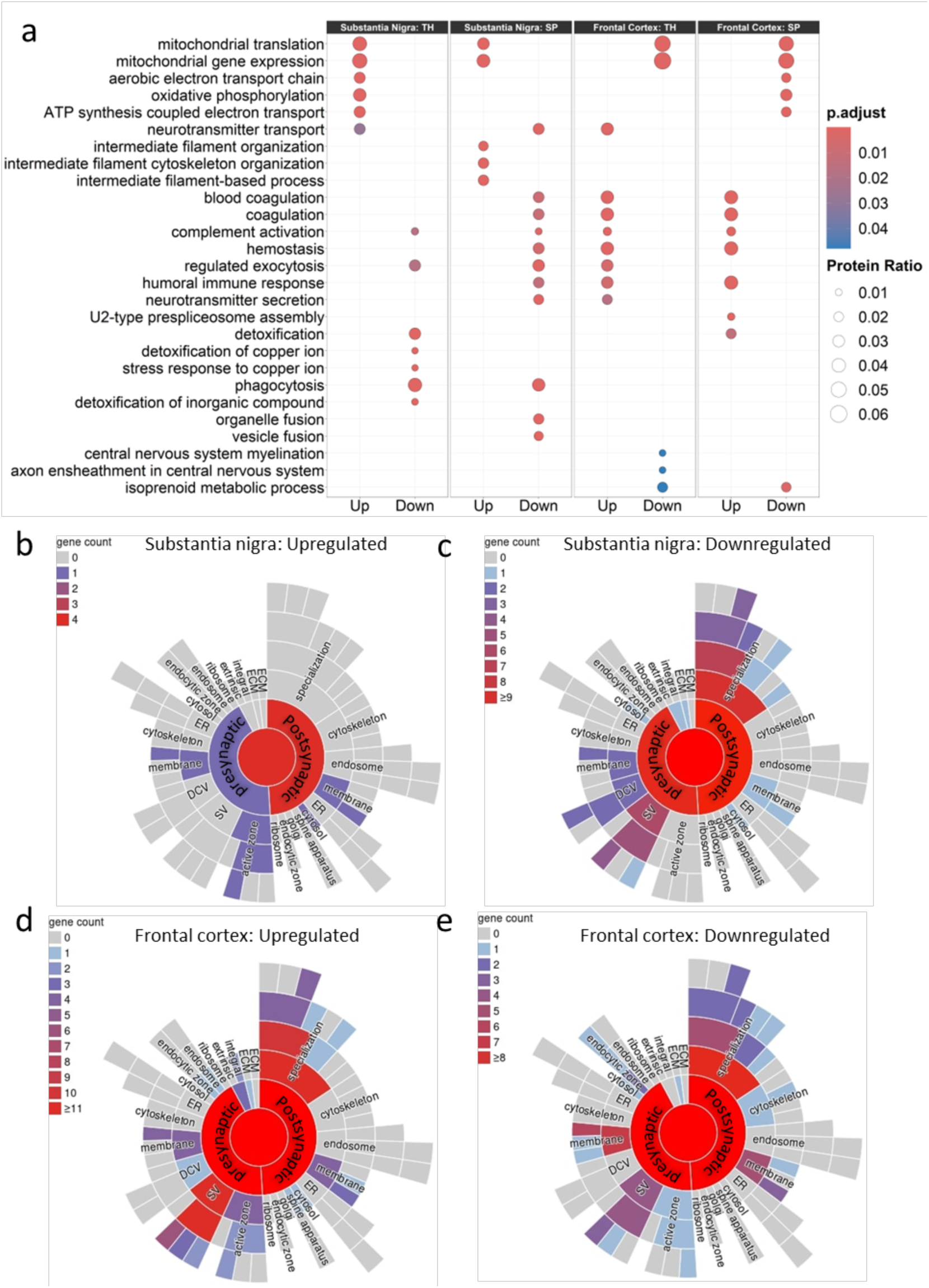
Proteomics analysis identifies dysregulated synaptic, metabolic and inflammatory pathways. Gene ontology enrichment analysis using the R package ‘clusterProfiler’ (setting: “biological processes”) was performed on DEPs highlighting that synaptic, metabolic and inflammatory pathways are altered in PSP. *p*-value adjusted using Benjamini-Hochberg correction for multiple comparisons. TH = total homogenate. SP = synaptoneurosome preparation (a). SynGO analysis was conducted on the synaptoneurosome preparations from the substantia nigra (b,c) and frontal cortex (d,e). In the substantia nigra there is a loss of pre- and post-synaptic proteins (b,c). In the frontal cortex, whilst there is also evidence for a reduction in some synaptic proteins (e) there is also evidence also some proteins that are increased (d).

### Synapses in living human brain tissue take up oligomeric tau derived from PSP brain

To study the effects of PSP-derived tau on living human brain, we developed a novel model system using living adult human neocortex donated as waste access tissue from surgical tumour resections (donor information in table 2), which was sliced into 350 μm sections, and challenged with post-mortem PSP brain tissue extract. Human brain slices were cultured for 72 hrs in 3 different conditions. Group 1 was cultured in 100% medium (control), Group 2 in 75% medium and 25% tau immunodepleted PSP brain soluble protein extract and Group 3 in 75% medium and 25% mock immunodepleted (thus tau containing) soluble PSP brain extract. Adjacent slices from each individual live tissue donor were treated with each of the 3 conditions providing an internal control for donor in a repeated measures design. After treatment, slices were collected for array tomography and western blot analyses. Array tomography ribbons were immunostained for GFAP-positive astrocytes, post-synapses (PSD95), phospho-tau Thr202, Ser205 (AT8) and oligomeric tau (T22). Slices cultured with the tau containing soluble PSP brain extract show an increase in the percentage of PSD95 puncta colocalising with T22 when compared to slices cultured in medium (β = 0.546, 95% CI [0.28, 075], *t*(9.56) = 6.07, *p* = 0.0004 ) or tau immunodepleted PSP brain extract (β = 0.423, 95% CI [0.19, 0.66], *t*(9.56) = 4.91, *p* = 0.002) (Fig. 6b). There is also a trend increase in the percentage of PSD95 puncta colocalising with AT8 in slices cultured with tau positive PSP brain extract, compared to slices cultured in medium alone (β = 0.138, 95% CI [0.003, 0.273], *t*(13.4) = 2.68, *p* = 0.046) or tau negative PSP sp-extract (β = 0.133, 95% CI [0.003, 0.270], *t*(13.4) = 2.57, *p* = 0.056) (Fig. 6c), however, the effect size is smaller compared to T22 and over 10-fold more post-synapses took up T22 than AT8 positive tau. These data suggest that PSP-derived tau can be taken up into living human post-synapses, and oligomeric tau has a greater propensity for synaptic uptake than AT8 positive tau species (phospho-tau Ser202, Thr205).

**Figure 6:**
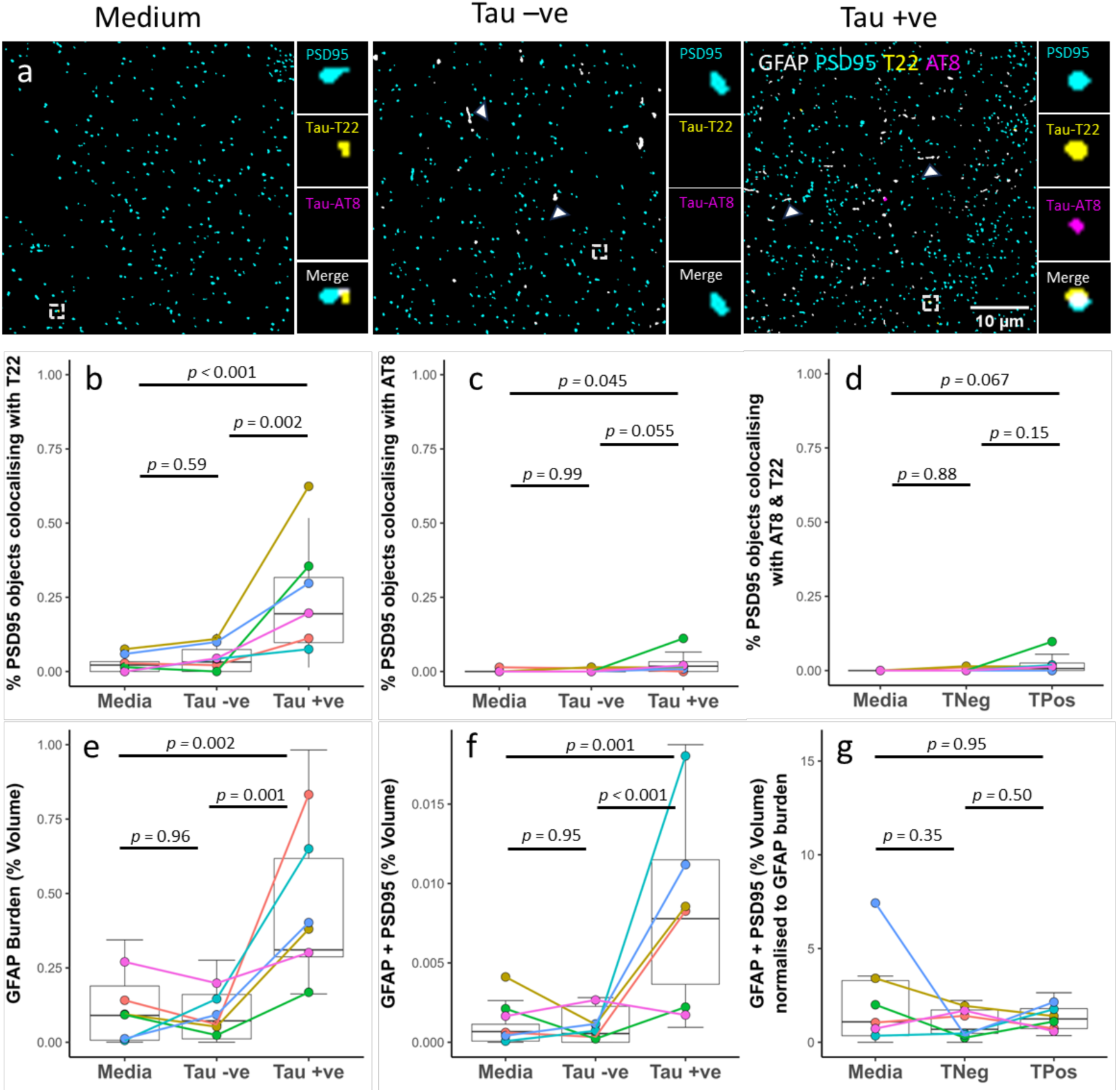
PSP derived tau induces astrogliosis, synaptic engulfment and is taken up into post-synapses in living human brain slices. **a)** Human organotypic brain slices were cultured for 72hrs in either medium only, tau –ve PSP sp-extract or tau +ve PSP sp-extract. Slices were processed for array tomography and immunostained for post-synapses (PSD95, cyan), oligomeric tau (T22, yellow) and phospho-tau Ser202, Thr205 (AT8, magenta) and astrocytes (GFAP, grey). Representative single 70nm segmented sections shown. Scale bar = 10um. Large boxes = 50um*50um. Small boxes = 2um*2um. Arrowheads show GFAP positive astrocyte processes. **b-g)** Quantification of colocalisation shows an increase in the percentage of PSD95 puncta with oligomeric tau (b), and a trend for an increase with AT8 (c) when cultured with the tau+ve PSP sp-extract. Slices cultured with the tau +ve sp-extract show an increase in the percentage volume of the 3D image stack occupied by GFAP (e) and the colocalisation of GFAP with PSD95 (f). However, when the colocalization between PSD95 and GFAP is normalised to the GFAP burden there is no difference between groups (g). Boxplots show quartiles and medians calculated from each image stack. Data points refer to case means. Colour = case.

**Table 2.** Details of cases used in human brain slice culture experiments (fig 6)

| Case | Sex | Age | Brain region | Tumour type | APOE |
| --- | --- | --- | --- | --- | --- |
| 01 | Male | 70s | Parietal Occipital | Glioblastoma | 3/3 |
| 02 | Female | 30s | Frontal | Glioblastoma | 2/4 |
| 03 | Female | 30s | Temporal | Glioma | 3/3 |
| 04 | Male | 50s | Frontal | Glioma | 3/3 |
| 05 | Female | 50s | Frontal | Metastases | 3/3 |
| 06 | Male | 50s | Temporal | Glioblastoma | 2/3 |

We then investigated the effects of PSP derived tau on astrocytes in human slice cultures. We found that there was a 3.4-fold increase in GFAP burden in slices cultured with tau containing PSP extract compared to medium alone (β = 0.35, 95% CI [0.14, 0.56], *t*(13) = 4.48, *p* = 0.002) and a 4.3-fold increase compared to tau immunodepleted PSP extract treatment (β = 0.37, 95% CI [0.16, 0.58], *t*(13) = 4.69, *p* = 0.001)(Fig. 6e). Further there was an 11.6-fold increase in colocalisation between PSD95 and GFAP compared to medium alone (β = 0.114, 95% CI [0.05, 0.18], *t*(12) = 4.75, *p* = 0.001) and a 14.2-fold increase compared to tau negative PSP sp-extract (β = 0.122, 95% CI [0.06, 0.19], *t*(12) = 5.00, *p* < 0.001)(Fig. 6f). When the amount of GFAP and PSD95 colocalisation was normalized to GFAP burden, there was no difference between treatments suggesting astrogliosis drives the increased synaptic ingestion (Fig. 6g).

## Discussion

Our study addresses important questions about how tau pathology spreads through the brain and how pathological tau leads to synapse degeneration in PSP. We demonstrate that: 1) tau accumulates within synaptic pairs in PSP both in an early and a late affected brain region; 2) pathological tau colocalises with synaptogyrin-3 in pre-synapes; 3) astrocytes show increased synaptic engulfment in PSP; and 4) that PSP-derived tau can be taken up into post-synapses and induce astrogliosis and augmented synaptic engulfment in living human brain slice cultures. Due to the scarcity of PSP post-mortem brain tissue suitably prepared for array tomography, our study is limited by our relatively small sample size and lack of power to observe any sex effects (36). However, in the available samples, we have been able to perform detailed investigation of synaptic changes in PSP.

Both the limits of resolution of light microscopy and the lack of living human brain model systems have previously limited the ability to directly study potential trans-synaptic spread of tau in PSP. Our experiments using a rare set of human post-mortem brain samples prepared for array tomography and a novel living human brain slice model (33,37) challenged with PSP brain extract, both support the idea that tau pathology spreads trans-synaptically in PSP. This is in line with much lower resolution human PET imaging data showing that tau pathology in PSP accumulates in functionally connected brain circuits (19), and with animal model data showing that human pathological tau spreads through brain regions via synaptic connections (12–17). In Alzheimer’s disease, we recently showed using array tomography and electron microscopy that tau likely spreads via synapses to occipital cortex, a late-affected brain region (20). Very recent data in a preprint using immuno-electron microscopy of human Alzheimer’s disease brain tissue, similarly supports trans-synaptic spread of phospho-tau through the Papez circuit including from the anterodorsal thalamic nucleus (38), implicating spread of tau pathology from subcortical regions. Our PSP data indicate that, similarly, tau may spread from subcortical regions (substantia nigra in our study) through circuits to the cortex. Our new data are important as they indicate that preventing pathological tau release from pre-synapses or preventing uptake by post-synapses could stop the spread of tau pathology through the brain in PSP. Since the spread of pathology is associated with symptom progression (3,4), this could be a promising disease-modifying approach.

Our finding that pathological tau accumulates inside synapses in PSP is also important as this likely contributes to synapse loss in this disease. Synaptic dysfunction is an early event associated with tau pathology in animal models of tauopathy, and synapses are lost in human tauopathies including PSP (39,40). Previous studies have demonstrated that tau and synaptogyrin-3 interact in mice and flies overexpressing human tau, causing clustering of synaptic vesicles, synapse loss, and cognitive impairment (9,10,27). Further, in human Alzheimer’s disease post-mortem brain, tau colocalises with synaptogyrin-3 (27). This study reveals that in PSP, tau colocalises in pre-synapses with synaptogyrin-3. Our data taken together with the previous animal data indicate that in primary tauopathies as well as AD, tau-synaptogyrin-3 interactions may be important for synaptic dysfunction and loss. Thus stopping this interaction is a potential therapeutic target to save synapses. Beyond synaptogyrin-3, our proteomics data highlights multiple changes in synaptic proteins. In substantia nigra which is affected early in the disease, we observe a loss of synaptic proteins. In frontal cortex, which is less affected by pathology much later, we see a reduction in some synaptic proteins but an increase in others hinting at a potential compensatory response earlier in the disease process. Of particular interest, we observe increased levels of syntaxin-6 in PSP frontal cortex in our proteomics analysis. Syntaxin-6 is a SNARE protein and several GWAS studies have identified a genetic link between the *STX6* gene and PSP (41–43). In a cell culture model, syntaxin-6 has been shown to facilitate tau secretion via binding tau at synaptic vesicles (44). The increase we observe in a late affected brain region support the idea that synatxin-6 could be important for synaptic tau spread in human PSP. Another potential mediator of synaptic spread identified is clusterin, which is increased in frontal cortex in PSP in our proteomics study. Clutsterin has been shown to promote tau seeding and propagation in a cellular model system via the formation of tau/clusterin complexes that enter recipient cells via endocytosis (45). Therefore, this upregulation of clusterin observed in PSP brain may be a mechanism driving post-synaptic uptake of pathological tau in this region.

Our study also indicates that astrocytes may play a role in synapse loss in PSP. In Alzheimer’s disease, we recently observed that both microglia and astrocytes contain synaptic protein with astrocytes engulfing more synapses than microglia (32). Here, we observed increased colocalization of synaptic proteins within astrocytes both in human post-mortem PSP tissue and in our human brain slices challenged with PSP-derived tau, indicating that this synaptic ingestion by astrocytes is not an end-stage phenomenon of the neurodegenerative disease. In our human slice experiments, immunodepleting tau from the PSP brain extract prevented astrogliosis and astrocyte engulfment of synapses, indicating that pathological tau in PSP brain specifically induces these phenotypes. Our proteomics results indicate there are molecular changes in astrocytes in PSP as we observe changes in levels of GFAP, YKL40/CHI3L1 and ICAM1 and several neuro-immune pathways. We further observe changes in microglial proteins and complement cascade proteins in PSP. This is in line with previous data implicating the complement pathway in the pathogenesis of PSP (43,46,47), and is of particular interest given the known role complement plays in mediating synaptic pruning (28,48–53). We also observed prominent dysregulation in proteins involved in mitochondrial function and metabolism in both the total brain homogenate and synaptic preparations from the substantia nigra and frontal cortex. This supports current evidence from laboratory, *in vivo* and post-mortem studies, that suggest mitochondrial energy metabolism is impaired in PSP (46,54–57).

In this study we applied sub-diffraction limit microscopy and biochemical enrichment of synapses to PSP brain to study pathological tau, synapses, and astrocytes and we further developed a novel model of living human brain tissue challenged with PSP-derived tau to examine tau uptake and astrocyte-synapse interactions. Based on our data, we conclude that tau pathology likely spreads at least in part via synapses in PSP and we reveal potential mechanisms leading from pathological tau to synapse degeneration. These data suggest that targeting synaptic tau may attenuate disease progression in PSP and should be considered as part of multi-modal therapy.

## Data Availability

Upon acceptance of the peer-reviewed version of this manuscript, all data spreadsheets and statistical analysis files will be available online as supplementary data. Images are available upon reasonable request. All custom software and scripts used in the study is available on GitHub at https://github.com/Spires-Jones-Lab

## Acknowledgements

We would like to thank patients and their families for providing tissue donations, without which this work would not be possible. We gratefully acknowledge the contributions of the Edinburgh Brain and Tissue Bank and Alzheimer’s Scotland Dementia Research Centre for coordinating post-mortem brain tissue donations, the NRS BioResource and Tissue Governance unit and EMERGE Research Nurse team for obtaining informed consent and surplus cortical tissue samples from NHS patients Edinburgh Neuroscience, the Genetics Core at the Edinburgh Clinical Research Facility University of Edinburgh for genotyping tissue taken from surgical resections Prof. Rakez Kayed for generously providing T22 antibody and Dr Paul Baxter, Dr Henner Koch, Dr Daniel Erskine, Dr Faye McLeod, Dr Crispin Jordan, Dr James Catterson and Dr Soraya Meftah for valuable discussions about our research. This work was supported by the UK Dementia Research Institute, which receives its funding from DRI Ltd, funded by the UK Medical Research Council, Alzheimer’s Society, and Alzheimer’s Research UK (grant code UKDRI-Edin005). The confocal microscope was generously funded by Alzheimer’s Research UK (ARUK-EG2016A-6) and a Wellcome Trust Institutional Strategic Support Fund at the University of Edinburgh. J.L. was funded by UCB Biopharma, as was the Oxford Nanoimager. M.H. and J.L. acknowledge funding from Dr. Jim Love. Dr Claire Durrant receives funding from Race Against Dementia (ARUK-RADF-2019a-001), The James Dyson Foundation, and the Alzheimer’s Society (581 (AS-PG-21-006)). Dr Robert McGeachan is funded by the Wellcome Trust, as part of the Edinburgh Clinical Academic Track for Veterinary Surgeons (225442/Z/22/Z). TSJ is a scientific advisory board member of Scottish Brain Sciences, Cognition Therapeutics, and Race Against Dementia and has consulted for Jay Therapeutics, and MT is an employee of Scottish Brain Sciences.

## Methods

### Human post-mortem samples and ethical approval

Post-mortem human brain tissue was acquired from the Edinburgh Brain Bank. The Edinburgh Brain Bank is a Medical Research Council funded facility with research ethics committee approval (16/ES/0084). The details of the post-mortem human cases (n=10 PSP, 13 control) are found in Table 1. As required by our Brain Bank ethical approvals, pseudoanonymized IDs are included in the table. PSP cases were included if they had a neuropathological diagnosis of PSP and a clinical diagnosis of a progressive neurodegenerative disorder. Of our 10 PSP cases, 6 had a clinical diagnosis of PSP (Steele-Richardson-Olszewski, subtype unknown), 3 had clinical diagnoses of unknown neurodegenerative conditions, and 1 had a diagnosis of Creutzfeldt-Jakob disease (which on pathology was found to be PSP not CJD). All 10 had neuropathologically confirmed PSP. Cases with significant co-pathologies were excluded. Control cases were matched for age (t-test, df=20.56, *t*=-1.34 *p* = 0.20), sex (x^2^ test, df=1, *χ^2^* = 0.25, *p* = 0.62) and post-mortem interval (PMI) (t-test, df=14.61, *t*=-1,78 *p* = 0.10). Exclusion criteria for control cases included known clinical neurological or psychiatric disease or a neuropathological report consistent with a different neurodegenerative disorder. The array tomography study size was limited to 7 per group by available tissue embedded for array tomography. Simulation-based power analysis (58) determined an n=7 would give a power = 0.78 to detect AT8 colocalisation with synaptophysin in PSP brain. Experiments were approved by the Edinburgh Brain Bank ethics committee, the Academic and Clinical Central Office for Research and Development (ACCORD) and the medical research ethics committee AMREC a joint office of the University of Edinburgh and National Health Service Lothian, approval number 15-HV-016. All data has been pseudoanonymized so personal identifying information cannot be accessed with these numbers.

### Array tomography: tissue processing and immunohistochemistry

Samples from the frontal cortex (Brodmann area 9 (BA9)) and substantia nigra (SN), were fixed and embedded for array tomography as described previously (Kay et al., 2013). Briefly, fresh tissue samples (∼1 cm3) were fixed in 4% paraformaldehyde for 3 hours, dehydrated in ethanol and incubated in LR White resin overnight at 4 °C. Individual samples were cured in LR White in gelatin capsules overnight at 53 °C. Samples from the frontal cortex (BA9) were cut into ribbons of ultrathin serial 70 nm sections using a histo Jumbo Diamond knife (Diatome) and an Ultracut (Leica). Ribbons were mounted on glass coverslips coated, dried on a slide warmer and outlined with hydrophobic pen. Ribbons were rehydrated in 50 mM glycine and blocked in a solution of 0.1% fish skin gelatin and 0.05% Tween in TBS for 45 minutes at room temperature. Ribbons were incubated with primary antibodies (Table 3) diluted in blocking solution overnight at 4°C. Ribbons were washed with TBS, and secondary antibodies (Table 3) diluted in blocking solution were applied for 45 minutes at room temperature. Ribbons were then washed with TBS. When used, direct label 488 synaptophysin diluted in blocking solution (1:200) was applied for 1 hour at room temperature, ribbons were then washed with TBS. Finally, ribbons were mounted on glass slides with ImmuMount. Images were obtained with a 63x 1.4 NA objective on an AxioImager (Zeiss) or Leica TCS confocal (Leica). Images from the same location in each serial section along a ribbon were aligned, thresholded, and parameters quantified using in house scripts in ImageJ, MATLAB (Mathworks), and Python. All software is freely available on GitHub (https://github.com/Spires-Jones-Lab). Saturation was minimized during image acquisition and only applied for figure visualization. 3D reconstructions were made with Imaris software (Bitplane). For more details, please see our methods video demonstrating this technique at https://doi.org/10.7488/ds/297. Image processing and analysis was performed blinded to the case ID.

**Table 3.** Antibody information.

| Antibody | Source | Identifier |
| --- | --- | --- |
| Rabbit anti-PSD95 | Cell Signaling Technology | Cat#3450; RRID:AB_2292883 |
| Guinea pig anti-PSD95 | Synaptic Systems | Cat#124 014;<br>RRID:AB_2619800 |
| Recombinant Alexa Fluor® 488 Anti-Synaptophysin antibody [YE269] | Abcam | Cat#ab196379;<br>RRID:AB_2922671 |
| Mouse anti-synaptophysin (SY38) | Abcam | Cat#ab8049;<br>RRID:AB_2198854 |
| Goat anti-synaptophysin | R and D Systems | Cat: #AF5555<br>RRID: AB_2198864 |
| Mouse anti-phosphorylated tau (AT8) | Thermo Fisher | Cat#MN1020;<br>RRID:AB_223647 |
| Rabbit anti-oligomeric tau (T22, serum) | Courtesy of Rakez Kayed | NA |
| Chicken anti-GFAP | Abcam | Cat#ab134436;<br>RRID: AB_2818977 |
| Rabbit anti-GAPDH | Abcam | Cat: #ab9485<br>RRID: AB_307275 |
| Mouse anti-synaptogyrin-3 | Santa Cruz | Cat: #sc-271046<br>RRID: AB_10611955 |
| Goat anti-human tau | (R&D systems | Cat: #AF3494<br>RRID: AB_573209 |
| Rabbit anti-histone (H3) | Abcam | Cat: #ab1791<br>RRID: AB_302613 |
| Goat anti-chicken Alexa Fluor® 405 | Thermo Fisher Scientific | Cat: #a48260<br>RRID: AB_2890271 |
| Donkey anti-guinea pig Alexa Fluor® 488 | Jackson ImmunoResearch Laboratories | Cat: #ab175658<br>RRID: AB_2340472 |
| Donkey anti-rabbit Alexa Fluor™ 594 | Invitrogen | Cat: #A-21207<br>RRID: AB_141637 |
| Donkey anti-mouse Alexa Fluor™ 594 | Invitrogen | Cat: #A-21203<br>RRID: AB_2535789 |
| Donkey anti-goat Alexa Fluor™ 594 | Invitrogen | Cat:#A-21447<br>RRID: AB_2535864 |
| Donkey anti-goat Alexa Fluor™ 405 | Abcam | Cat:# ab175664<br>RRID: AB_2313502 |
| Goat anti-mouse IgG1 Alexa Fluor™ 647 | Invitrogen | Cat: #A21240<br>RRID: AB_2535809 |
| Donkey anti-rabbit IRDye 800CW | Li-Cor Biosciences | Cat:# 925-32213<br>RRID: AB_2715510 |
| Donkey anti-mouse IRDye 680RD | Li-Cor Biosciences | Cat# 925-68072<br>RRID: AB_2814912 |

### Microscopy and Image Analysis

Experimenters were blinded to case information during image processing and analysis. Images were acquired using Zen software using a 63x oil immersion objective (1.4 numerical aperture) on a Zeiss AxioImager Z2 with a CoolSnap digital camera or using a Leica SP8 TCS confocal microscope. For array tomography, wwo regions of interest (ROIs) were chosen in the gray matter of BA9 or pars compacta SN, and imaged in the same location on 15-30 sequential sections. Imaging parameters were kept the same throughout each experiment, and an image was taken of the negative control in each session to ensure there was not non-specific staining.

Individual images from each ROI were combined into a 3D image stack and a median background filter was applied in Image J with custom batch macros. Using custom MATLAB scripts, image stacks were aligned using rigid registration. Each channel was segmented using an auto-local thresholding algorithm to binarize images and remove any objects present in only a single section (from secondary antibody noise). Segmented images were run through custom MATLAB and Python scripts to determine object density and colocalization between channels. Objects were considered colocalized if at least 25% of the 3D volume overlapped. For pre- and post-synaptic objects to be considered a synaptic pair, the distance between the centre of each object had to be ≤0.5 μm. 3D reconstructions were made with Imaris software (Bitplane). Saturation was minimized during image acquisition and only applied for figure visualization. All custom software scripts are available on GitHub https://github.com/Spires-Jones-Lab. For more details, please see our methods video demonstrating this technique at https://doi.org/10.7488/ds/297.

### Soluble protein extraction from PSP brain and tau immunodepletion

5.3 grams of PSP frontal cortex (BA6/8) was thawed on ice, finely diced and homogenized in 30mL 1X artificial cerebral spinal fluid (aCSF, pH 7.4) supplemented with 3 complete mini EDTA-free protease inhibitor cocktail tablet (Roche, 11836170001) using a Dounce homogenizer. Homogenized tissue was transferred to 15 mL protein LoBind® tubes (Eppendorf, 0030122216), placed on a roller for 30 minutes at 4°C, and then centrifuged at 2000g and 4°C for 10 minutes to remove insoluble debris. The supernatant was centrifuged again for 110 minutes at 200,000g and 4°C. The resulting supernatant, which here we will call the soluble protein extract (sp-extract) was dialysed in aCSF for 72 hours (Slide-A-Lyzer G2 Dialysis Cassettes) to remove salts, impurities and drugs the donor took. The aCSF was replaced every 24 hours. Following dialysis, the sp-extract was pooled and divided in two. The two portions of sp-extract were incubated with Protein A agarose (PrA) beads (30 μL / ml of sample) (ThermoFisher Scientific, 20334) and either a total tau antibody Tau13 (mouse IgG1, 1:50, biolegend, #MMs-520R) to create the “tau negative” (TNeg) fraction or an anti-GFP antibody (mouse IgG1, 1:50, DSHB, DSHB-GFP-8H11) to create the “tau positive” (TPos) fraction, overnight at 4 °C on a rotating mixer. Then the solutions were centrifuged at 2500 g for 5 minutes, the supernatant was collected into fresh protein LoBind® tubes and incubated again with a fresh PrA beads / antibody solution for a total of 4 times. Finally, the tau positive and tau negative supernatant was collected into fresh protein LoBind® Eppendorf’s and stored at -80 °C. The concentration of tau in each of the sp-extract was quantified by sandwich enzyme-linked immunoassay (ELISA) according to the manufacturer’s instructions (ThermoFisher Scientific, #KHB0041). The total tau concentration in the tau negative sp-extract = 0.013 μg/ml and in the tau positive sp-extract = 11.14 μg/ml. Sp-extracts were aliquoted in protein low bind tubes and stored at -80°C. All work involving human tissue was conducted in a dedicated fume hood, all work involving tissue or aCSF was performed at 4°C, and all contaminated consumables were disinfected in Virkon broad-spectrum virucidal disinfectant overnight before being disposed of in appropriate waste systems.

### Tissue and ethical approval for Human Organotypic Brain Slice Cultures

Human brain slice cultures (HBSCs) were made using surplus, peritumour cortical access tissue surgically resected from patients undergoing glioblastoma debulking surgery as previously described (33,59). Use of living resected human tissue has been approved by AMREC and Lothian NRS Bioresource (Spires-Jones and Durrant, REC number: 15/ES/0094, IRAS number: 165488, Bioresource No SR1319). Additional Caldicott Guardian approval (CRD19080) has been obtained to receive data about the sex, age (in years), brain region the tissue was taken from and the clinical reason for surgery. The informed consent of patients was obtained using the Lothian NRS Bioresource Consent Form. Case information for the patients used in this study is outlined in Table 2.

### Preparation and Maintenance of Human Brain Slice Cultures

To generate HBSCs, surplus peritumoural human brain tissue was collected at surgery into ice cold and continuously oxygenated artificial cerebrospinal fluid (aCSF) (Supplementary material, Table 1). The peritumour cortical tissue was then embedded in 4% agar and the resulting block superglued (Loctite to the vibratome stage. The block of tissue was sectioned using a Leica VT1200S vibratome into 300 µm thick sections. The tissue was further dissected so that each slice contained all the cortical layers and a small amount of white matter for orientation (Figure 2C). Slices were then placed into a wash buffer composed of continuously oxygenated Hanks Balanced Salt Solution (HBSS) and HEPES (20 mM) (305 mOsm, pH 7.3) for 15 minutes at room temperature. Slices were then plated on 0.4-μm pore membranes (Millipore: PICM0RG50) sitting on top of 750 μl of a second wash solution (BrainPhys Neuronal Culture Medium (StemCell Technologies) (96%), N2 (1X), B27 (1X), hBDNF (40 ng/ml), hGDNF (30 ng/ml), Wnt7a (30 ng/ml), ascorbic acid (200 nM), dibutyryl cAMP (1 mM), laminin (1 ug/ml), penicillin-streptomycin (1%), nystatin (3 units/ml) and HEPES (20mM)). Slice cultures were kept in the second wash medium in an incubator at 37 °C with 5% CO2 for 1-2 hours, after which the medium was aspirated and replaced with 1 of 3 different solutions. Group 1 was cultured in 100% maintenance medium (same as “second wash solution” above, but without HEPES). Group 2 was cultured 75% maintenance medium and 25% tau negative PSP brain sp-extract (final total tau concentration <0.003 μg/ml). Group 3 was cultured in 75% maintenance medium and 25% tau positive PSP brain sp-extract (final total tau concentration = 2.789 μg/ml). The aCSF, wash solutions and maintenance medium were passed through a 0.22μm-filter prior to use to ensure sterility. To prevent the removal of protein aggregates, the PSP sp-extracts were not filtered, but we did not encounter contamination issues. After 72 hours the slices were processed for array tomography and immunostained as detailed above, the fixation time was reduced to 90 minutes to account for a reduced thickness of tissue compared to post-mortem samples. The antibody information is listed in table 3.

### Proteomics

Total brain homogenate and biochemically isolated synapses were prepared for proteomics as previously published(60). Briefly, 300-500 mg of PSP or control frontal cortex or substantia nigra was homogenised using a Dounce homogeniser with 1ml of buffer (2 5mM HEPES, 120 mM NaCl, 5 mM KCl, 1 mM MgCl2, 2 mM CaCl2, protease inhibitors (Merck, 11836170001) and phosphatase inhibitors (Merck, 524629-1SET), made up in sterile water). The Dounce homogeniser and buffer were kept ice-cold throughout the procedure to reduce degradation. We allowed 10 passes for full homogenisation but minimise cellular disruption that would lead to an impure final fraction. Once homogenised, the homogenate was aspirated in a 1ml syringe and passed through a pre-soaked 80 μm filter (Merck, NY8002500) to remove debris and yielded the total brain homogenate (TH). At this stage a sample of TH was snap-frozen on dry ice for proteomics analysis. The remaining TH sample was passed through a pre-soaked 5-micron filter (Merck, SLSV025LS) followed by centrifugation at 2000 g for 5 minutes to yield the synaptoneurosome (SN) pellet. The supernatant was discarded and the pellet was washed with buffer, and centrifuged a second time at 2000 g for 5 minutes to ensure purity. The supernatant was discarded and the pellets were snap frozen on dry ice and stored at - 80°C. Both the 80 μm and 5 μm filters were pre-soaked with 1 ml of Buffer A to maximise yield.

Samples we diluted five-fold with Tris-HCl buffer pH 7.6 (100 mM Tris-HCl, 4% SDS, and Protease inhibitor cocktail EDTA-free [ThermoFisher Scientific, 78447]), and homogenised thoroughly with a pipette tip. The samples were centrifuged for 20 minutes at 17,000 G at 4 °C. The supernatant was aliquoted into fresh protein LoBind® Eppendorf’s (Protein LoBind) and stored at -80 °C. Micro BCA™ Protein Assay kit (ThermoFisher Scientific, 23235) was used to quantify protein levels, following manufacturer’s instructions. Briefly, working solution was made fresh right before required, consisting of 50% Buffer A, 48% Buffer B and 2% Buffer C, all provided in kit. Briefly, 0.5μl of sample was added to 200μl of working solution. Albumin was provided in the kit for determining a standard curve of the following protein concentration: 2 μg/ml, 4 μg/ml, 6 μg/ml, 8 μg/ml, 10 μg/ml, 20 μg/ml and 40 μg/ml. Absorbance values were obtained using spectrophotometry at 562 nm with working solution used as the blank value to calibrate the machine.

Peptide samples were resuspended in 0.1% formic acid prior to loading into vials for Mass Spectrometry analysis. Samples were acquired on a Thermo Exploris 480 (Thermo) connected in-line with a Ultimate 3000 UPLC (Thermo). 1 µg of peptides were loaded in 1 µL onto a 5 µm, 100 µm x 2cm nanoViper C18 trap column (Thermo). Peptides were separated using a 2µm, 75 µm x 50cm C18 reversed phase Easy-spray analytical column over 135 minutes at a flow rate of 300 nl/min. A linear gradient from 3% to 35% was used from water with 0.1% formic acid to acetonitrile with 0.1% formic acid. MS data was acquired in data-independent mode using a 45 variable m/z window method.

### Western Blot

After protein extraction, each sample was made-up to 15 μg of protein in 15 μl of de-ionised water as calculated by the micro BCA™ and diluted in half with Laemmli buffer (2x stock) (S3401-10VL). In each well, 15 μl of sample were loaded in 4-12% Bis-Tris gels (ThermoFisher Scientific, NP0323BOX). Each gel was run with 5 μl of molecular weight marker (Licor, 928-40000) in the first well. were washed with de-ionised water and diluted NuPAGE buffer (20x stock) to remove bits of broken gel and residual acrylamide. Western blot chambers were filled with diluted NuPAGE buffer (ThermoFisher Scientific, NP0002). Gels were run at 80 V for 5 minutes followed by 120V for 1.5 hours. Gels were washed in 20% ethanol for 10 minutes prior to transferring using the iBlot 2 Dry Blotting System IB21001 as per manufacturer’s instructions. Pre-packed transfer stacks containing a PVDF membrane (ThermoFisher Scientific, IB24002) were assembled, and samples were transferred for 8.5 minutes at 25 V. After transferring we used the Li-Cor Revert 700 total protein stain kit (Li-Cor Biosciences: 926-11010), for western blot normalisation, as per the manufacturer’s instructions. Briefly, membranes were incubated with Revert 700 total protein stain for 5 minutes, washed twice for 30 seconds in Revert 700 wash solution, imaged using a Li-Cor Odyssey Fc machine, and then de-stained for 7 minutes in the Revert de-staining solution. Membranes were then blocked for 1 hour using PBS Intercept Blocking Buffer (Li-Cor Biosciences: 927-70001). Primary antibodies were diluted in PBS Intercept Blocking Buffer with 0.1% Tween-20 and incubated with membranes overnight at room temperature (Synaptophysin 1:1000 (abcam, ab8049); PSD95 1:1000 (Cell signalling technologies, D27E11); Histone 1:2000 (abcam, ab1791). Membranes were washed three times for 5 minutes with PBS-Tween, then incubated in darkness for 2 hours with secondary antibodies IRDye 800CW Donkey anti-rabbit (Li-Cor Biosciences: 925-32213) and IRDye 680RD Donkey anti-mouse (Li-Cor Biosciences: 925-680RD), both at 1:5000 concentration. Membranes were washed 3X in PBS-Tween, 1X in PBS and then imaged using a Li-Cor Odyssey Fc machine. Synaptoneurosome preparations were assessed for enrichment for synaptophysin or PSD95 and exclusion of histones (Supplementary fig. 3a-c). Preparations that did not show enrichment for synaptic proteins were repeated.

### Electron Microscopy

Synaptoneurosome pellets were fixed in 4% Paraformaldehyde, 2.5% Glutaraldehyde, 0.2% Picric acid in 0.1 M phosphate buffer, pH 7.4 for 2 hours at room temperature plus overnight at 4 °C, post-fixed in 1% Osmium tetroxide for 30 minutes, washed in 0.1 M PB, boiled distilled water, and 50% ethanol, incubated in 1% uranyl acetate in 70% ethanol, dehydrated through 15 min steps in a graded series of ethanol then propylene oxide, 50% propylene oxide/50% Durcupan resin, then 100% Durcupan resin overnight in a Leica EM TP processor. Samples were baked in Durcupan resin in agar capsules overnight at 56C, cut on an Ultracut microtome (Leica) with a Histo Jumbo diamond knife (Diatome) into 50 nm sections which were mounted on nickel mesh grids. Electron micrographs were captured on a Zeiss Gemini 360 scanning electron microscope with an annular STEM detector.

### Statistical analyses

Statistical analysis was performed using RStudio with R version 4.3.1. The analysis and visualization of the data were facilitated by utilizing the large language model ChatGPT, which was employed to assist in refining R code. The majority of statistical analyses used linear mixed effects models (LMEM)(‘lme4’ R package), as this allowed us to test if diagnosis or treatment group impacted our variable of interest while controlling for potentially confounding variables, such as age and post-mortem interval (PMI), and including random effects to account for repeated measures. The decision to ultimately include potentially confounding fixed and random effects in the model, and prevent overfitting of the model, was done by inspecting Akaike Information Criterion (AIC) and Bayesian Information Criterion (BIC) to assess which model best fits the data. Linear mixed effect models assume linearity, normal distribution of residuals and homogeneity of variance. Linearity was assessed by plotting model residuals against predictors, normality of residuals was checked with a QQ-plot, and homogeneity of variance checked by plotting residuals against fitted values. If the model did not meet the assumptions, data was transformed using the method that was best transformed each individual model to fit the assumptions. Data transformations included square root, log, arcsine square root, and Tukey transformation. Post-hoc testing was conducted for pairwise comparisons, estimated marginal means and 95% confidence intervals (’emmeans’ package), with p-values adjusted using Tukey correction for multiple comparisons. Effect sizes are displayed as ‘β’ with 95% confidence intervals (CI). When data was transformed to meet the model assumptions then the reported effect sizes are computed on the back transformed data. Degrees of freedom were calculated using Kenwood-Roger approximation. Correlations used Pearson’s or Spearman’s methods, dependent on whether data met assumptions for parametric testing or not. Statistical details for each analyses can be found in the results text. Mass spectrometry .raw files were processed using DIA-NN (version 1.8.1 doi: 10.1038/s41592-019-0638-x) using the ‘library free’ method. A human uniprot FASTA (downloaded on 01/09/2023) was used as the reference database for library generation. The default settings were used except for Heuristic protein inference. Protein inference was set to “Protein names” and the double-pass mode for the neural network classifier was used. Resultant protein abundance matrices were then processed using R (version 4.1.3). Protein abundance values were normalised using the quantile normalisation as part of the PreProcessCore package. Missing values were imputed using the ImpSeqRob function as part of the rrcovNA package using the default parameters. Prior to statistical comparison using the Limma package. Comparisons were made using the “eBayes” method. Differentially expressed proteins (DEPs) were defined as proteins that exhibited more than a 20% alteration in protein levels (fold change < 0.8 or > 1.2) and a *p*- value < 0.05. The choice of a 20% change as a threshold was motivated by the consideration that such alterations likely represent biologically meaningful shifts in protein levels. DEPs were analyzed utilizing Gene Ontology (GO) enrichment (’clusterProfiler’ R package) (34) and the synaptic gene ontology tool, SynGO (35) with a *p*-value cut off = 0.05, *q*-value cut off = 0.05 and *p*-values adjustment method set to Benjamini-Hochberg correction for multiple comparisons.

### Data Sharing

Upon acceptance for publication, all spreadsheets and statistical analysis files will be shared on Edinburgh Datashare https://datashare.ed.ac.uk/handle/10283/3076. Proteomics data will be shared through the PRIDE repository. Raw images and data available from the lead authors upon reasonable request.

---

**Supplementary figure 1:**
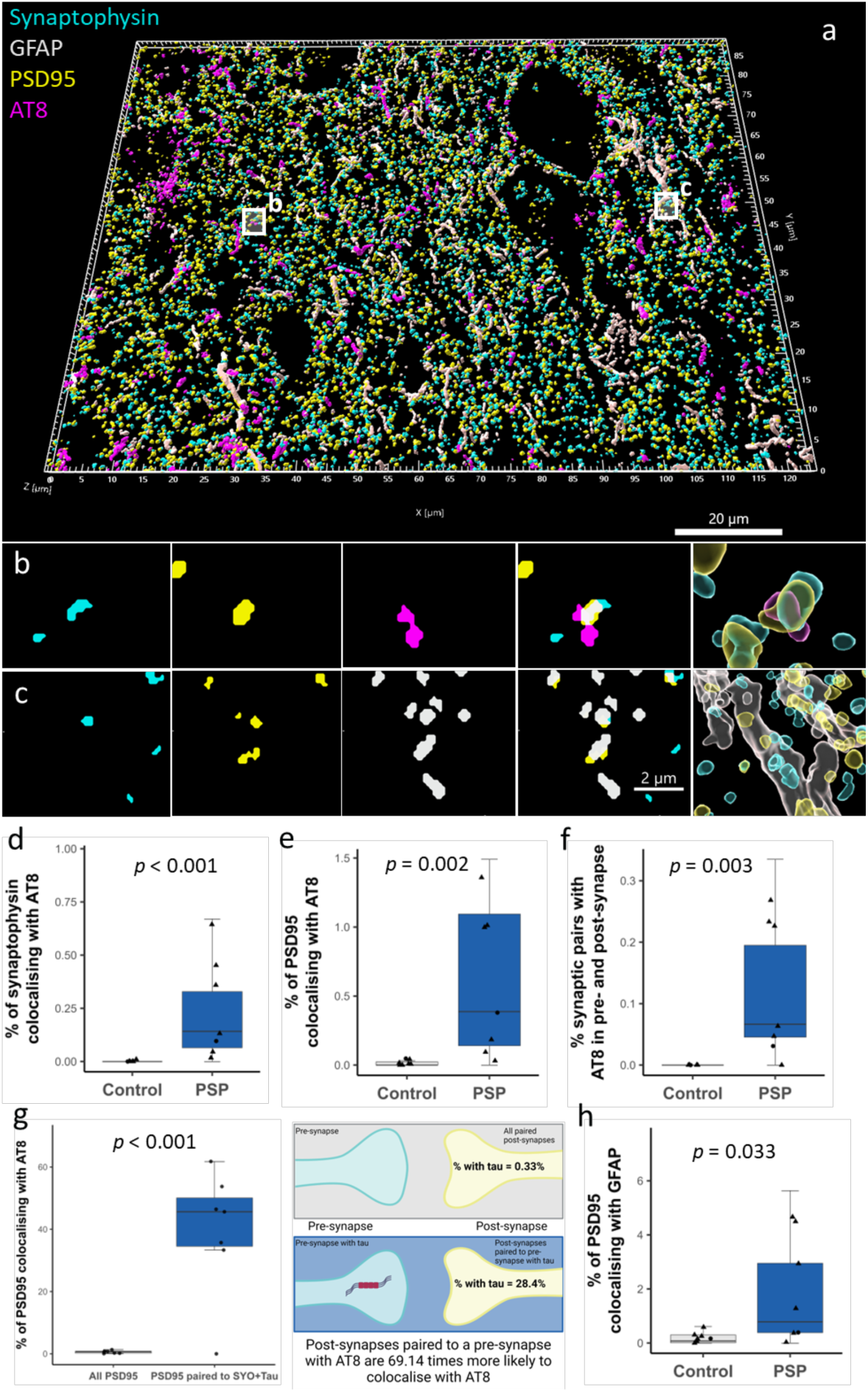
Evidence for the transsynaptic spread of phospho-tau 202/205 and augmented post-synaptic engulfment by astrocytes in PSP frontal cortex. **a-c)** A 3D reconstruction of PSP frontal cortex, immunostained for pre-synapses (synaptophysin, cyan), post-synapses (PSD95, yellow), tau (AT8, magenta) and astrocytes (GFAP, grey) (a). Tau (AT8) is seen to accumulate in synaptic pairs (b) and pre-synapses and post-synapses are seen within GFAP positive astrocyte processes (c) 3D reconstructions in a and far right panels of b and c made using IMARIS. **d-h)** Quantification shows that in PSP frontal cortex there is an increase in the percentage of pre-synapse (d), post-synapses (e) and synaptic pairs (f) colocalising with tau (AT8). Post-synapses that are paired to a pre-synapses with tau (AT8) are around 69.14 times more likely to colocalise with tau (AT8) than all paired post-synapses, regardless of whether the adjacent pre-synapse also colocalises with tau (AT8). There is also an increased colocalisation between post-synapses (PSD95) and astrocytes (GFAP) (h). Boxplots show quartiles and medians calculated from each image stack. Data points refer to case means (females, circles; males, triangles). Schematic created with biorender.com.

**Supplementary figure 2:**
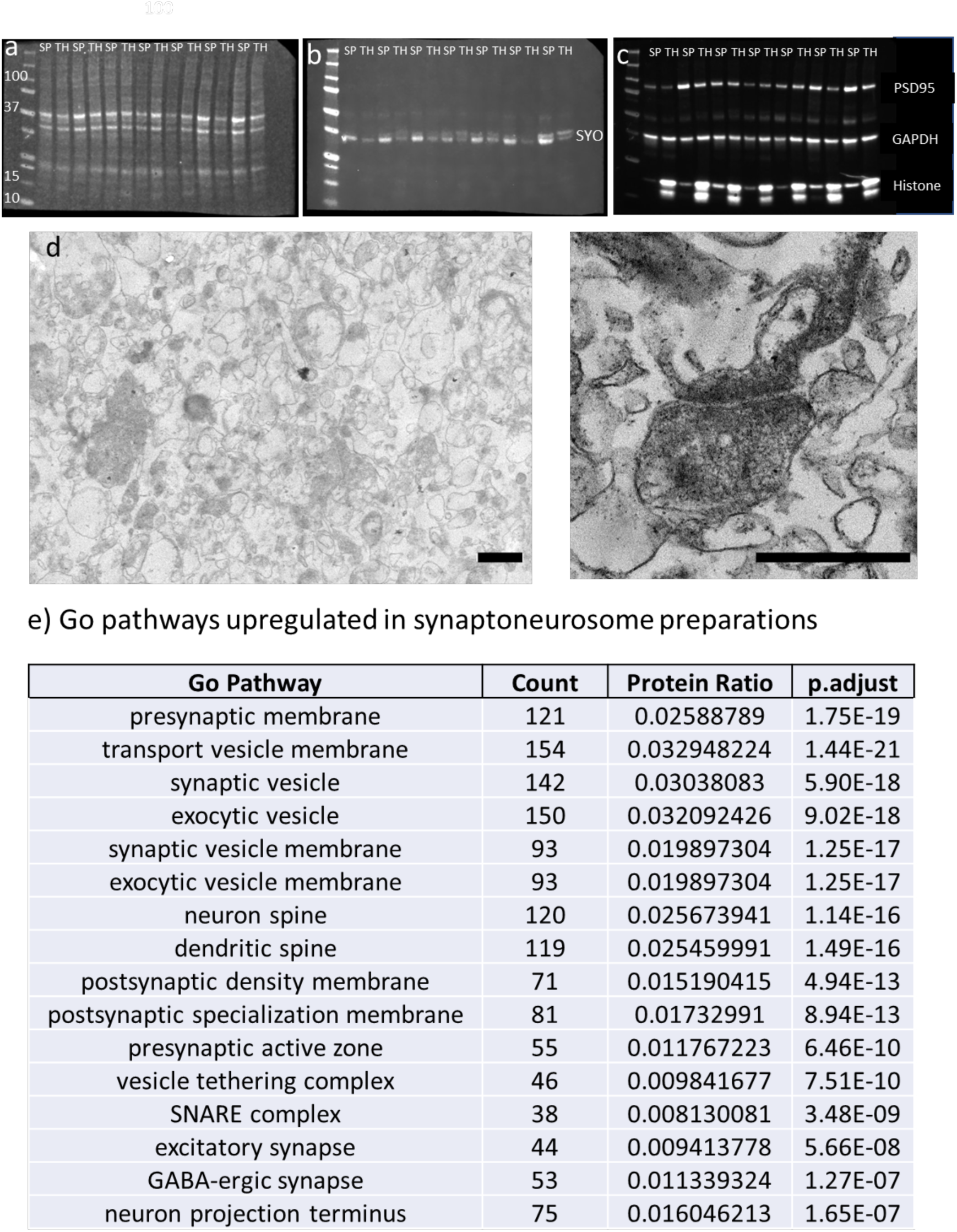
Validation of enrichment of synaptic proteins. **a-c)** Western blot analysis reveals enriched synaptic proteins (synaptophysin and PSD95) and decreased nuclear protein (histones h3) in synaptoneurosome preparations. Membrane was probed with REVERT total protein stain (a), synaptophysin (SYO)(b), and PSD95, Histones H3 and GAPDH (c). SP = Synaptoneurosome preparation. TH = total homogenate. **d)** Electron microscopy of synaptoneurosome preparations show exclusion of cell bodies and enrichment of synapses. Scale bar left image = 1μm, Scale bar right image = 0.5μm **e)** Gene ontology enrichment analysis using the R package ‘clusterProfiler’, setting = “cellular compartment”, was performed on DEPs between total homogenate and synaptoneurosome preparations and revealed an enrichment of synaptic proteins.

**Supplemental figure 3:**
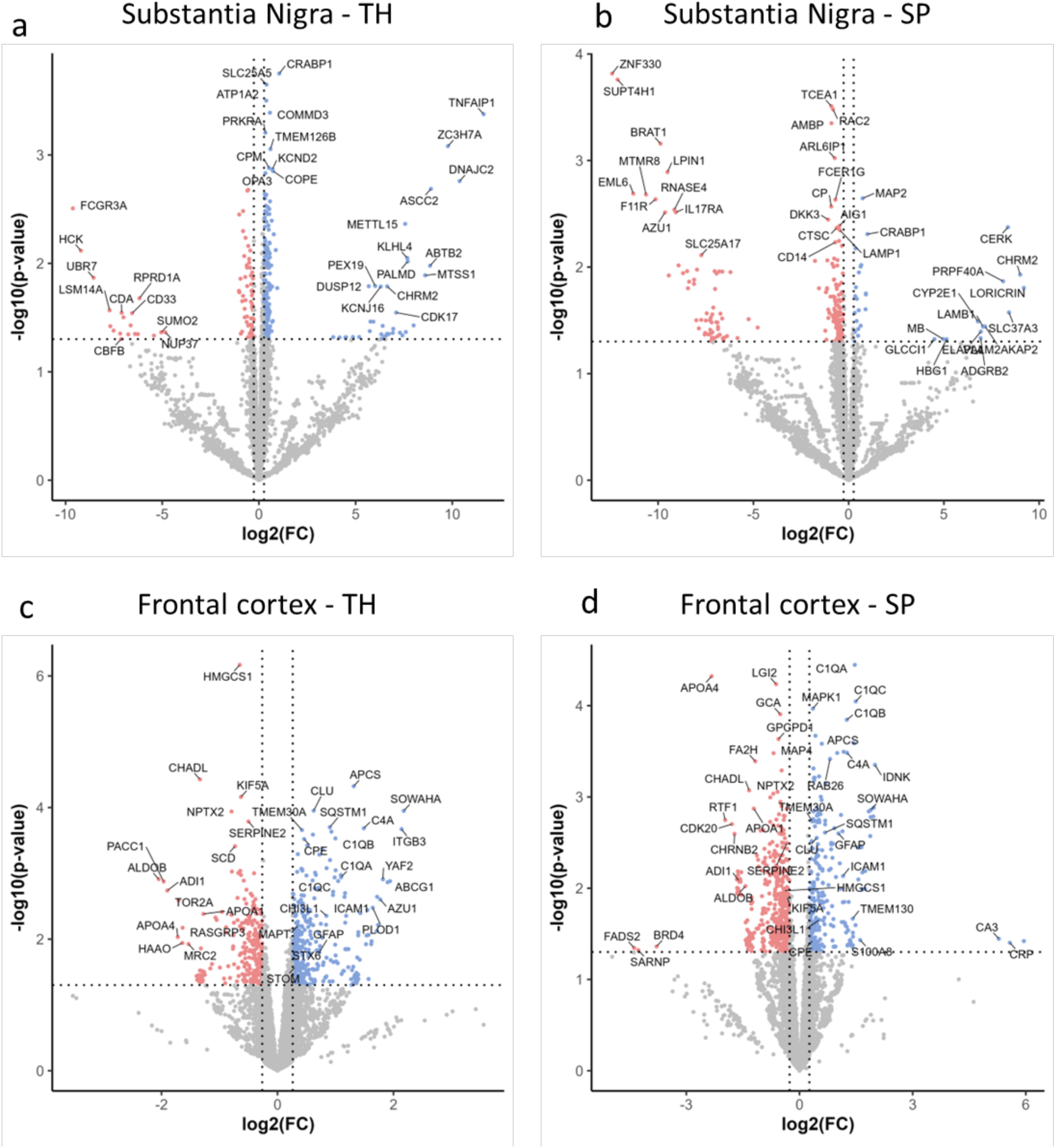
Volcano plots showing differentially expressed proteins in PSP substantia nigra and frontal cortex. **a-d)** Volcano plots highlighting the results of proteomic analysis comparing PSP and control substantia nigra total homogenate (a), substantia nigra synaptoneurosome (b), frontal cortex total homogenate (c) and frontal cortex synaptoneurosome (d). Differentially expressed proteins (DEPs) were defined as *p*-value <0.05 and 20% change in protein level. Down regulated proteins shown in red. Upregulated proteins shown in blue. Proteins that were not differentially expressed are highlighted in grey. TH = total homogenate, SP = synaptoneurosome preparation.

